# Hospital Discharge Prediction Using Machine Learning

**DOI:** 10.1101/2024.06.20.24309256

**Authors:** Joaquim Oristrell, Anna Pascual, Pere Millet, Guillermo R. Lázaro, Anna Benavent

**Author notes:** **CORRESPONDING AUTHOR:** Joaquim Oristrell, MD, PhD, Emeritus Researcher. Parc Taulí Hospital Universitari. Institut d’Investigació i Innovació Parc Taulí (I3PT-CERCA). Universitat Autònoma de Barcelona. 08208 Sabadell (Barcelona). Catalonia. Mail.

## Abstract

**OBJECTIVE:** Reliable hospital discharge predictions still remain an unmet need. In this study, we aimed to forecast daily hospital discharges by ward, until seven days ahead, using machine learning methods.

**METHODS:** We analyzed all (n=67308) hospital admissions proceeding from the Emergency department in a University Hospital, from January-2018 to August-2023. Several train-test splits were defined simulating a prospective, weekly acquisition of data on new admissions. First, we trained Light Gradient Boosting Machines (LGBM) and Multilayer Perceptron (MLP) models to generate predictions on length of stay (LOS) for each admission. Based on predicted LOS, timeseries were built and predictions on daily hospital discharges, by ward, seven days into the future, were created employing diverse forecasting techniques. Mean absolute error (MAE) between predicted and observed discharges was used to measure the accuracy of predictions. Discharge predictions were also categorized as successful if they did not exceed by 10% the mean number of hospital daily discharges.

**RESULTS:** LGBM slightly outperformed MLP in 25 weekly LOS predictions (MAE 4.7±0.7 vs 4.9±0.7 days, p<0.001). The best techniques to forecast, seven days ahead, the daily number of hospital discharges were obtained using Prophet (MAE 5.0, R^2^=0.85), LGBM (MAE 5.2, R^2^=0.85), seasonal ARIMA (MAE 5.5, R^2^=0.81) and Temporal Fusion Transformer (TFT)(MAE 5.7, R^2^=0.83). After categorizing the predictions, LGBM, Prophet, seasonal ARIMA and TFT reached successful predictions in 82.3%, 81.1%, 77.7% and 77.1% of days, respectively.

**CONCLUSIONS:** Successful predictions of daily hospital discharges, seven days ahead, were obtained combining LOS predictions using LGBM and timeseries forecasting techniques.

**Lay abstract:** Currently, most public hospitals in western countries have close to full occupancy for significant periods of time. Under these conditions, it is common for emergency admissions to be delayed, which causes significant patient discomfort and can negatively impact their quality of care. Predicting the daily number of hospital discharges would enable hospital administrators to implement measures to prevent hospital overcrowding.

In this study, we used several artificial intelligence methods to predict, seven days in advance, the number of daily hospital discharges, obtaining successful predictions in more than 80% of the days that were analyzed.

In conclusion, we have shown that available machine learning methods offer new and valuable options to predict hospital discharges, until seven days in advance, with high efficiency and reliability.

**HIGHLIGHTS:**

1. Accurate predictions of hospital discharges could enable optimization of patient flow management within hospitals.
2. Emerging machine learning and time-series forecasting methods present novel avenues for refining hospital discharge predictions.
3. In this study, we integrated length of stay predictions using Light Gradient Boosting Machines with several time-series forecasting techniques to produce daily hospital discharge forecasts.
4. Through the combined used of these methodologies, we were able to obtain successful predictions on more than 80% of the days.

## Introduction

Currently, most public hospitals in western countries have close to full occupancy for significant periods of time. Under these conditions, it is common for emergency admissions to be delayed, which causes significant patient discomfort and can negatively impact their quality of care.

Predicting the daily number of hospital discharges would enable hospital administrators to implement measures to prevent hospital overcrowding [1]. Although many studies have analyzed the factors that impact on hospital stays in specific services or populations [2–6], forecasting the overall daily number of hospital ward discharges is complex and has received less attention [7].

In recent years, some experiences using machine learning methods to predict hospital discharges have been published, some of them based on administrative data with few or no clinical covariates [8–10], and others being modeled to generate discharge forecasts only 24h in advance [11–13], which is a very short time lapse for hospital managers to be able to adapt hospital resources to the predicted healthcare demands.

In this study we aimed to predict the number of daily discharges by ward, seven days into the future, in a large university hospital. First of all, using clinical variables, we predicted the individual length of stay (LOS) with two different machine learning algorithms. Secondly, the number of hospital and ward daily discharges was predicted based on individual LOS predictions and calendar-related variables using a tree-based and timeseries forecasting methods.

## Material and Methods

### Design

We retrospectively analyzed the electronic medical records of the patients discharged from the Corporació Sanitaria Parc Taulí (CSPT). Our institution is a 714-bed public university hospital serving a population of 394,000 in Barcelona county (Catalonia).

We included all the patients ≥18 years being admitted from the Emergency department (ED) and discharged from our hospital from 1^st^ January 2018 until 31^st^ August 2023. The patients admitted in 2020 were excluded from the analysis due to the uncommon hospital dynamics as a result of the COVID’s pandemic. Programmed admissions, and patients admitted to Pediatrics, Psychiatry or Rehabilitation services were excluded.

To generate forecasts, a prospective inclusion of patients was simulated. All hospitalized patients until 25 index dates (every Monday from March 6 until August 21, 2023) were analyzed. First of all, for each patient, a LOS prediction was made based on variables collected at the time of admission (see section “Patient-related variables” described below). Secondly, aggregating the patients admitted before each index date by their predicted day of discharge, a timeseries dataframe with the predicted number of discharges per day, by departments and wards, was obtained.

Since new, unknown, admissions occur between each index date and the day to be predicted, a second model was developed, using a timeseries approach, that included the predicted number of discharges per day of the previous model as well as other temporal and autoregressive variables (see section “Calendar-related variables” described below), combining an estimate of discharges that will occur for known patients as well as discharges for patients yet to be admitted.

### Variables used for the analysis

*Patient-related variables.* All patient-related variables were collected in the ED, before hospital admission. These included the age, gender, ED diagnoses and comorbidities, medical procedures performed or requested from the ED, prescription of broad-spectrum antibiotics (Anatomical Therapeutic Chemical codes J01DH or J01CR05), number of ED visits in the last 6 months, weekday at admission, and time elapsed from ED admission until hospital admission. Finally, the clinical service, hospital ward, physician on charge, and LOS were also collected.

Principal diagnoses were encoded using ICD-10-CM and grouped into 1195 categories according to pathophysiology or 1211 categories according to organ involvement. Principal diagnoses and comorbidities were also classified into 159 common syndromes. Finally, a comorbid index was developed using the B coefficients of comorbidities that significantly impacted on LOS in a linear regression model (Supplementary Table 1). ED procedures were also classified using ICD-10-PCS codes and 53 variables identifying the most common emergency procedures were generated.

*Calendar-related variables:* the timeseries dataframe with predicted discharges by day was completed with variables identifying weekdays, non-working days, and their respective lagged variables. Sine and cosine Fourier’s terms were also included as independent variables to capture weekly or monthly patterns in hospital discharges using the *numpy.fft* function of the NumPy package, version 1.23.5 [14].

### Prediction of length of stay (LOS)

We used Light Gradient Boosting Machines (LGBM) and Multilayer Perceptron (MLP) to predict the LOS for all admissions occurring until 25 index dates. LGBM is a tree-based, open access machine learning method developed by Microsoft [15,16], while MLP is a network of fully connected neurons with a non-linear activation function [17]. For both methodologies, we employed patient-related variables collected at the ED before hospital admission. Mean absolute error (MAE) was used as the metric applied to the loss function. Further details on LGBM and MLP methods applied are described in the Supplementary Methods section.

### Predictions of daily number of hospital discharges

Weekly forecasts on daily number of hospital discharges were generated with LGBM and compared with six timeseries forecasting methods: Seasonal ARIMA with exogenous variables (SARIMAX), Prophet, Long-Short term memory (LSTM), Temporal Fusion Transformer (TFT), Neural Hierarchical Interpolation for timeseries forecasting (N_HiTS), and Temporal Convolutional Network (TCN).

The SARIMAX model is a statistical timeseries forecasting technique that extends the traditional Autoregressive Integrated Moving Average (ARIMA) model to account for seasonality and additional covariables [18]. Prophet is a non-linear, additive regression model developed by Facebook that captures the linear trend and uses Fourier’s terms to capture the seasonality of the timeseries. Holiday or other categorical effects can also be added as dummy variables [19,20]. LSTM is a recurrent neural network used for natural language processing and timeseries forecasting [21]. TFT is a neural network developed by Google that allows to generate timeseries forecasts adding attention mechanisms to LSTM-type encoders [22]. N_HiTS is a recently described neural network architecture for timeseries forecasting based on multiple interconnected blocks of MLP networks [23]. Finally, TCN has been developed for timeseries forecasting using several stacked, one-dimensional, convolutional networks, feeding only from past observations to forecast future timesteps [24].

#### Building the timeseries

After obtaining the LGBM-predicted length of stay for each episode, the predicted date of discharge was calculated by adding the predicted length of stay to the corresponding date of admission. Grouping the episodes by the predicted date of discharge, hospital departments, wards and gender resulted in department and ward-specific timeseries dataframes, each one with predicted and real discharges by date. These timeseries dataframes were completed with known calendar-related variables and splitted into train and test datasets. The real number of discharges in each test dataset were set to missing before running the predictions. All this process was repeated for each index date, from 2023-03-06 until 2023-08-21, in order to obtain 25 train-test datasets and weekly predictions on daily discharges. For hospital discharge predictions using LGBM, we used the whole study period. However, due to the exclusion of year 2020 admissions, we only included data from January 2021 to August 2023 for all timeseries forecasting methods. Further details on LGBM and timeseries methods applied to predict the daily number of discharges are described in the Supplementary Methods section.

#### Comparing the prediction methods and classification of the predictions

Mean absolute error (MAE) and median absolute errors (MdAE) were the metrics applied to compare the different machine learning methods. In addition, Akaike Information Criterion [25] was also calculated for predictions performed in three index dates (6^th^ March, 8^th^ May and 10^th^ July 2023), and their mean values were used to compare between the most efficient methodologies.

Finally, as an excess of real discharges over predicted ones usually does not raise significant concerns for patient flow, we categorized daily predictions as unsuccessful if the predicted discharges exceeded by 10% the mean number of daily discharges; otherwise, they were considered successful.

### Statistical analysis

Continuous variables are reported as mean and standard deviation and categorical variables are summarized by frequencies and percentages. Associations between continuous variables were analyzed using Pearson’s correlation. Differences on continuous variables between groups were assessed using Student’s t test if univariate or linear regression for multivariate analysis. These analyses were performed using the Python’s scipy version 1.10.1 [26] and statsmodels version 0.14.0 [27] packages.

### Ethics

All data were treated anonymously in order for this study to comply with the provisions of Spanish and European laws on Protection of Personal Data. The study was approved by the ethics committee of the CSPT.

## Results

A total of 67308 hospitalization episodes proceeding from the Emergency department (ED) took place in the Medical (n=35935), Surgical (n=14156), Gynecological-Obstetrical (n=8533) or Short-stay (n=7677) departments of the CSPT during the period of study. Additionally, 1007 patients were discharged from the Intensive Care Unit (57.1% due to exitus, and 24.3% being transferred to other hospitals).

Mean age of the admitted patients was 65.3 (Standard deviation [SD] 20.7) years and 51.8% were women. The length of hospital stay showed an exponential distribution with a mean length of stay of 8.1 days (SD 11.4; range 0-355; median 4.0) (Supplementary Figure 1). Mean number of diagnoses established in the ED was 1.4 (SD 0.9), and mean number of past comorbidities was 4.2 (SD 3.9). A more detailed description of the characteristics of the episodes of hospitalization included in this study is depicted in Table 1.

**Table 1.** Characteristics of the hospitalization episodes included in the study.

|  | Total | Medical department | Surgical department | Gynecological-Obstetrical department |  | Short-stay department |
| --- | --- | --- | --- | --- | --- | --- |
|  |  |  |  | *Obstetrics service* | *Gynecology service* |  |
| Admissions, No. | 67308 | 35935 | 14156 | 7786 | 747 | 7677 |
| Female, No. (%) | 34876 (51.8) | 16004 (44.5) | 6231 (44.0) | 7786 (100) | 747 (100) | 3758 (49.0) |
| Age, mean(SD) | 65.3 (20.7) | 72.7 (15.7) | 62,8 (20.0) | 31.9 (5.8) | 39.1 (13.4) | 71.8 (15.9) |
| LOS<sup>1</sup>, mean(SD) | 8.1 (11.4) | 10.2 (12.3) | 9.5 (12.8) | 2.2 (2.3) | 2.6 (4.4) | 1.8 (1.7) |
| N° Diagnostics, mean(SD) | 1.4 (0.9) | 1.6 (1.0) | 1.2 (0.5) | 1.1 (0.4) | 1.2 (0.5) | 1.4 (0.7) |
| N° Previous Diagnostics, mean(SD) | 4.2 (3.9) | 5.5 (3.9) | 2.8 (3.2) | 0.5 (0.9) | 1.2 (1.9) | 5.0 (3.6) |
| N° Procedures, mean(SD) | 0.5 (1.1) | 0.6 (1.2) | 0.6 (1.1) | 0.1 (0.5) | 0.4 (0.8) | 0.6 (1.2) |
| Comorbidity index<sup>2</sup>, mean(SD) | 0.86 (1.53) | 1.24 (1.82) | 0.51 (1.20) | 0.02 (0.17) | 0.12 (0.44) | 0.84 (1.42) |
| Patients with chronic conditions, No.(%) | 13739 (20.4) | 9935 (27.6) | 1,892 (13.3) | 2 (0.03) | 4 (0.5) | 1801 (23.5) |
| Patients with advanced chronic conditions<sup>3</sup>, No.(%) | 1,614 (2.4) | 1380 (3.8) | 121 (0.9) | 0 (0) | 0 (0) | 111 (1.4) |
| Patients with low Barthel index, No.(%) | 6893 (10.2) | 5634 (13.8) | 847 (6.0) | 0 (0) | 0 (0) | 383 (5.0) |
| Patients on broad_spectrum antibiotics, No.(%) | 5079 (7.5) | 3424 (9.5) | 1395 (9.9) | 3 (0.04) | 9 (1.2) | 131 (1.7) |
| Number of previous ED attendances<sup>4</sup>, mean(SD) | 2.0 (1.6) | 2.0 (1.5) | 1.8 (1.4) | 2.7 (2.1) | 2.3 (1.8) | 2.0 (1.8) |

<sup>1</sup>LOS: length of stay; <sup>2</sup>Comorbidity index (see text); <sup>3</sup>Patients with chronic conditions with <1 year of expected life expectancy; <sup>4</sup>Number of previous Emergency department visits during the last 6 months.

### Prediction of length of stay (LOS)

Weekly LOS predictions for new admission episodes occurring from 6^th^ March until 27^th^ August 2023 are shown in Supplementary Table 2. LOS predictions using LGBM slightly outperformed MLP, with MAE values of 4.7 (SD 0.7) for 25 weekly LGBM predictions versus 4.9 (SD 0.7) for MLP, t-test p<0.001) (Supplementary Table 2).

Due to these results, the predicted number of daily discharges inferred from LGBM LOS predictions was used, in addition to calendar-derived variables, for the final prediction of daily hospital discharges.

Feature importance in LGBM LOS predictions is shown in Supplementary Figure 2. Median stay by ward and service, and mean or median stay by diagnosis (ungrouped and grouped), age, time spent in the ED before admission, main procedures, use of broad spectrum antibiotics, and the comorbidity index were the most important variables to predict LOS.

Using univariate and multivariate linear regression analysis (Supplementary Table 3) we observed that higher errors in LGBM LOS predictions were associated with higher comorbidity indexes, admission on Medical or Surgical departments (instead of Obstetrics or Short stay wards), use of broad spectrum antibiotics, or being diagnosed with a disease with longer mean LOS.

### Prediction of hospital daily discharges

Mean number of daily discharges was 38.9 (SD 17.6) for all admitted patients proceeding from the ED. Mean daily discharges for medical, surgical, obstetrical and short-stay departments were 21.0 (SD 12.9), 8.3 (4.4), 4.5 (SD 2.1) and 4.6 (SD 2.5), respectively. Daily discharges in our hospital showed a strong weekly seasonal pattern, with important fluctuations between weekends and working days, more evident for medical and surgical departments than for other services (Supplementary Figure 3).

Forecasts on hospital daily discharges were generated using LGBM and diverse timeseries methodologies. Mean and SD of MAE for 25 weekly predictions of whole hospital discharges was 5.0 (SD 1.7) using Prophet; 5.2 (SD 1.6) for LGBM; 5.5 (SD 1.7) with SARIMAX; 5.7 (SD 1.7) for TFT; 6.2 (SD 6.0) for LSTM; 6.5 (SD 2.8) for TCN; and 7.7 (SD 6.7) discharges for N_HiTS. Similarly, Pearson’s correlations between predicted and observed whole hospital discharges were R^2^=0.85 for LGBM and Prophet, 0.83 for TFT, 0.81 for SARIMAX, 0.76 with LSTM, 0.72 for TCN and 0.66 with N_HiTS. Mean MAE of all predictions on discharges, by forecasting method, department and ward are detailed in Table 2.

**Table 2.** Accuracy of different machine learning methods to predict the daily number of hospital discharges, by departments and wards.

|  | Prophet | LGBM | SARIMAX | TFT | LSTM | TCN | N_HITS | No. daily discharges,<br>mean |
| --- | --- | --- | --- | --- | --- | --- | --- | --- |
| **AIC** | 1.8 | 77.8 | 1.8 | 105.8 | 101.8 | 73.6 | 101.8 |  |
| **All departments. MAE, mean** | 5.0 | 5.2 | 5.5 | 5.7 | 6.2 | 6.5 | 7.7 | 38.9 |
| All departments. Females. MAE, mean | 3.2 | 4.0 | 3.3 | 3.2 | 4.0 | 3.5 | 4.6 | 20.2 |
| All departments. Males. MAE, mean | 3.6 | 3.8 | 3.6 | 4.2 | 4.4 | 4.7 | 5.6 | 18.7 |
| **Medical department. MAE, mean** | 4.2 | 3.8 | 4.0 | 4.5 | 4.8 | 5.2 | 5.7 | 21.0 |
| **Surgical department. MAE, mean** | 2.5 | 2.7 | 2.4 | 2.1 | 2.7 | 3.0 | 2.8 | 8.3 |
| **Obstetrical department. MAE, mean** | 1.6 | 1.9 | 1.7 | 1.6 | 1.7 | 1.4 | 1.6 | 4.5 |
| **Short stay department. MAE, mean** | 1.6 | 1.9 | 1.6 | 1.6 | 1.5 | 1.6 | 1.5 | 4.6 |
| **Ward 1, MAE, mean** | 1.7 | 1.6 | 1.7 | 1.6 | 1.8 | 1.7 | 1.9 | 4.8 |
| **Ward2, MAE, mean** | 1.3 | 1.3 | 1.4 | 1.4 | 1.5 | 1.7 | 1.5 | 4.0 |
| **Ward3, MAE, mean** | 1.4 | 1.2 | 1.5 | 1.3 | 1.4 | 1.6 | 1.5 | 3.9 |
| **Ward4, MAE, mean** | 1.3 | 1.2 | 1.4 | 1.3 | 1.4 | 2.2 | 1.4 | 2.9 |
| **Ward5, MAE, mean** | 0.7 | 0.7 | 1.0 | 0.7 | 0.8 | 0.6 | 0.7 | 1.6 |
| **Ward6, MAE, mean** | 1.4 | 1.1 | 1.6 | 1.2 | 1.7 | 1.3 | 1.5 | 3.6 |
| **Ward7, MAE, mean** | 1.5 | 1.4 | 1.5 | 1.5 | 2.1 | 2.1 | 1.5 | 3.2 |
| **Ward8, MAE, mean** | 2.7 | 3.1 | 2.3 | 3.1 | 2.8 | 3.3 | 2.2 | 4.4 |
| **Ward9, MAE, mean** | 1.6 | 2.1 | 1.8 | 1.7 | 1.9 | 1.9 | 1.7 | 5.3 |
| **Ward10, MAE, mean** | 1.6 | 1.9 | 1.6 | 1.6 | 1.5 | 1.6 | 1.5 | 4.6 |

AIC: Akaike Information Criterion. SARIMAX: Seasonal ARIMA with exogenous variables. LGBM: Light Gradient Boosting Machines; TFT: Temporal Fusion Transformer; LSTM: Long Short Term Memory; TCN: Temporal Convolutional Network; N\_HITS: Neural Hierarchical Interpolation for timeseries forecasting. MAE (mean absolute error) values are the mean MAE of 25 weekly predictions of discharges, from March 6 to August 27.

The best results for hospital discharge prediction were obtained using Prophet, LGBM, SARIMAX or TFT (Table 2, Figure 1). Prophet and SARIMAX offered a lower Akaike Information Criterion compared to LGBM and TFT since they reached similar predictions with lower number of variables. LGBM obtained the best predictions in medical wards, while SARIMAX and Prophet generated slightly better results in the surgical department (Table 2, Supplementary Fig 4).

**Figure 1.**
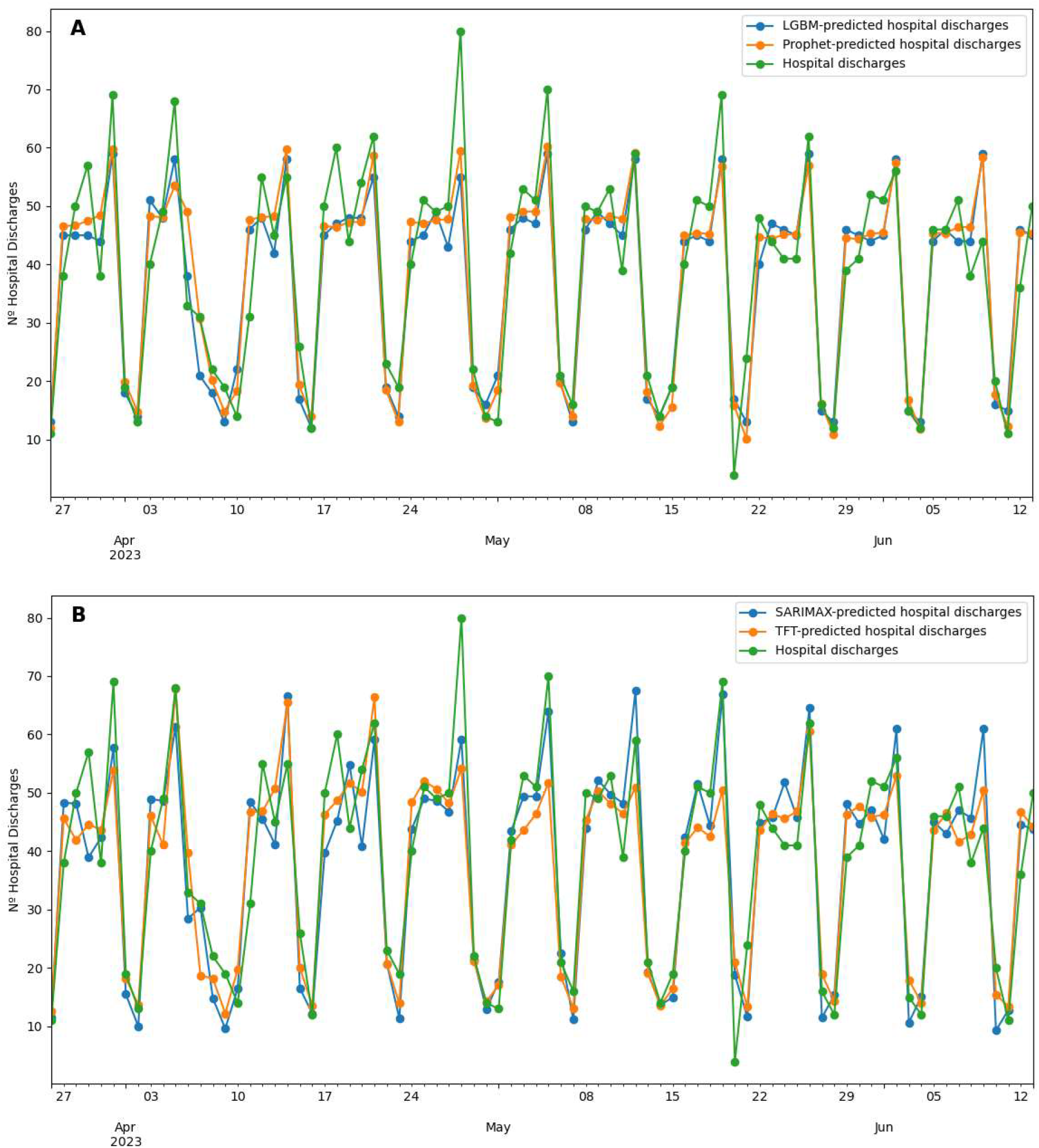
Concatenated weekly predictions of all hospital discharges. A: Light Gradient Boosting Machines and Prophet. B: SARIMAX and Temporal Fusion Transformer.

The most relevant variables for predicting daily hospital discharges using LGBM, as revealed by feature importance analysis, were the number of LOS-predicted discharges, lagged-values of daily admissions and discharges, transformed Fourier’s sinus and cosinus variables, and a category identifying the non-working days (Supplementary Fig 5).

### Proportion of dates with a successful prediction

Finally, as already mentioned, predictions for the whole hospital were classified as unsuccessful if they exceeded by 10% the mean number of daily discharges. With a mean number of 38.4 daily discharges we considered an unsuccessful prediction if the number of predicted discharges, for the whole hospital, exceeded in >4 the number of observed discharges.

According with this definition, we observed that 144 (82.3%), 142 (81.1%), 136 (77.7%) and 135 (77.1%) of the 175 dates between 6^th^ March and 27^th^ August 2023 were successfully predicted with LGBM, Prophet, SARIMAX or TFT respectively (Figure 2).

**Figure 2.**
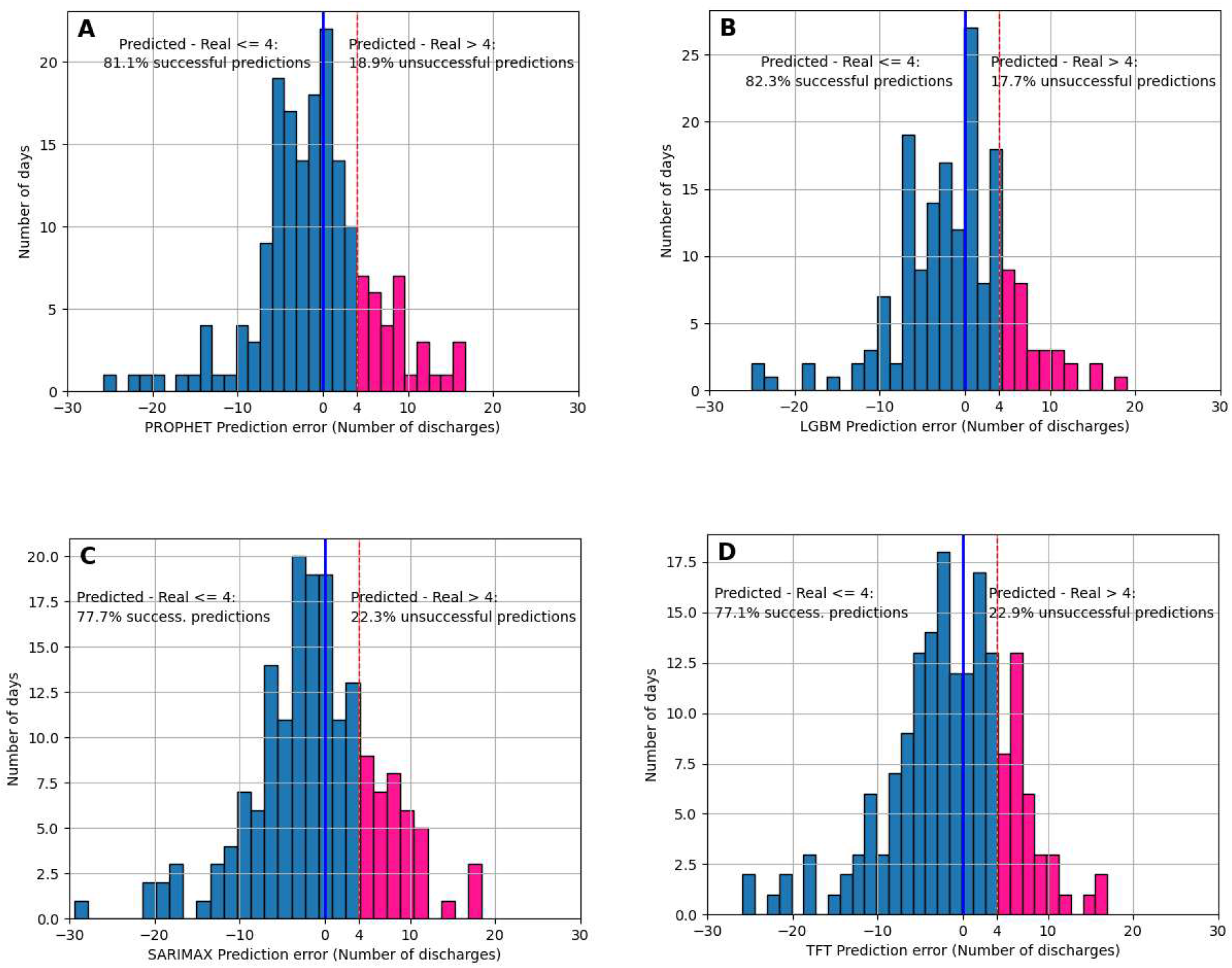
Categorization of discharge predictions as successful or unsuccessful (see criteria on the text) on 175 consecutive days. A. Prophet’s predictions. B. Light Gradient Boosting Machines predictions. C. SARIMAX predictions. D. Temporal Fusion Transformer predictions.

## Discussion

In this study we have combined individual length of stay predictions with temporal series forecasting to predict the hospital discharges by department and wards, seven days in the future from 25 index dates. This approach outperforms previous studies, most of them aimed to predict daily hospital discharges based on the length of stay (LOS) of each particular admission episode without considering trends or seasonalities [28–31], or predicting daily discharges using timeseries forecasting methods without considering individual clinical variables that impact on LOS [8,10].

Our study is retrospective. However, we managed to make 25 weekly predictions, each one based on new admissions along the last week, simulating a prospective collection of data and generating weekly predictions for a period of five months. Our results show that it is feasible to obtain a significant estimation of the number of daily discharges by ward, up to seven days in advance, with a single weekly acquisition of data on new admissions, making it easy to be applied in many settings. We only analyzed hospital admissions proceeding from the ED. It is conceivable that the inclusion of programmed admissions (i.e. programmed surgery), which often have a more predictable hospital stay [12] would probably improve our predictions.

First of all we tried to compare LGBM, a tree-based algorithm, with a MLP neural network to predict individual LOS. In our assays, a correctly tuned LGBM to prevent overfitting offered better results than MLP, at higher speed, and with no need of scaling input data. We are not aware of previous attemps to use LGBM to predict LOS, although other boosting techniques have been tried and showed better results than classical tree-based methods like random forest [32]. In our opinion, the superiority of LGBM could be explained by its known high efficiency in handling large datasets with many categorical variables and sparse data [15]. Among the limitations for LOS predictions we should mention that we did not include some variables that could probably impact on better LOS predictions, like the social status (i.e. patients living alone, poverty,..), laboratory values, or variables on drug therapies other than broad-spectrum antibiotics. In addition, we only analyzed the diagnoses and procedures performed or requested from the ED. It is probable that incorporating more precise diagnoses achieved during hospitalization, or new procedures performed after hospital admission, would probably improve the predictions. In any case, since we intended to predict the discharges several days in advance, unlike some previous studies [12], we did not find convenient to take into account the diagnoses established at discharge.

Secondly, after performing individual LOS predictions, we compared LGBM and diverse timeseries forecasting methods to predict hospital daily discharges. In the past, linear regression models [3] and classical timeseries analysis with ARIMA, seasonal ARIMA, ARMA or exponential smoothing have already been applied to forecast hospital patient flow with little success [9,33]. SARIMAX was previously used to predict hospital admissions and occupancy [10], but it has not been applied, to our knowledge, to forecast hospital discharges.

In recent years, new and efficient computational tools have appeared for timeseries forecasting that have now been assayed in this study. Facebook’s Prophet, described in 2017, has already been assayed to predict hospital discharges using holidays as the unique covariate [8]. Using the predicted number of daily discharges (derived from predicted LOS) as an additional covariate, we have been able to increase Prophet’s predictability.

In our opinion, although very good predictions could be obtained with Prophet or SARIMAX, both methods are rather simple prediction tools, where forecasts are mainly built on past values of the target variable, with few covariates, so that they could generate misleading predictions in changing scenarios like the opening or closing of new hospital wards, or if significant changes in inhospital patient composition do occur.

LGBM also offered very good predictions. In the past, other tree-based methods, like random forest or XGBoost, have been used in classification tasks, to identify hospitalized patients that will probably leave the hospital 24 hours in advance [12,34]. We are not aware that LGBM, or any other boosting technique, have been used in regression tasks, as we did, to predict the daily number of hospital discharges. LGBM is a flexible method, that can be adjusted with covariates, rapid to execute and interpretable.

We also tried four different timeseries neural network architectures (LSTM, N_HiTS, TCN and TFT). We are not aware of previous attemps to use, neither of them, to predict hospital discharges. Among these methods, TFT clearly reached the best predictions, very close to those obtained with LGBM, Prophet or SARIMAX, but with significantly higher computational costs.

Finally, we categorized the predictions as successful if they did not exceed by 10% the mean number of daily discharges, in a similar way as other authors have applied to classify predictions on hospital admissions [10]. Using this approach, successful predictions in more than 80% of the dates were obtained with LGBM and Prophet.

In conclusion, we have shown that available machine learning methodologies and timeseries forecasting techniques offer new and valuable options to predict hospital discharges by ward or department, until seven days in advance, with high efficiency and reliability. We have also shown that a two-phase forecast, initially predicting LOS using LGBM and subsequently forecasting the number of daily discharges applying LGBM or Prophet, yielded the best results. These methods should enable the departmental directors and hospital managers to better adjust hospital resources to future healthcare needs.

## Data Availability

Data produced in the present study are available upon reasonable request to the authors

## SUPPLEMENTARY MATERIAL

### 1.- Supplementary Methods

#### 1.- PREDICTING THE LENGTH OF STAY (LOS)

##### a) Prediction of LOS using Light Gradient Boosting Machines (LGBM)

Patient-related variables were used to build LGBM models using the Python’s lightgbm package, version 3.3.5 [1,2]. Training datasets comprised all patients admitted from 1^st^ January 2018 until 78 days before each index date, while the test datasets involved the patients admitted during the last 77 days before each index date. The length of the test periods was chosen empirically after trying different proportions of train/test periods. Limits to the number of leaves, and a minimum number of episodes by leave were established to prevent LGBM trees to overfit. Finally, all models were validated by means of 5-fold cross-validation on the training dataset.

##### b) Prediction of LOS using Multilayer Perceptron (MLP)

A MLP neural network was built with the TensorFlow package, version 2.13.0 [3], using the same covariables and test periods as for LGBM. For MLP each training period was splitted into a train and a validation dataset, the latter including the admissions of the last 4 months of the training period. Different network architectures, number of epochs and learning rates were assayed, plotting the validation loss curves to control overfitting. The final model was a three-layered MLP, with an input layer capturing 376 different variables, a hidden layer with 4 fully connected neurons and a dense fully connected output layer.

#### 2.- PREDICTING THE DAILY NUMBER OF HOSPITAL/DEPARTMENT/WARD DISCHARGES

##### Hospital discharge predictions with LGBM

For hospital discharge prediction with LGBM we also used the Python’s lightgbm package, version 3.3.5. The train-test split was set to 120 days before each index date, extending the test period until seven days ahead from each corresponding index date.

Seven LGBM models, one for each day of the week were built to predict the number of discharges by hospital department and ward, seven days ahead from each index date. Mean absolute error (MAE) was the metric applied to the loss function. Limits to the number of leaves, and a minimum number of days by leave were established to prevent LGBM models to overfit.

##### Hospital discharge predictions using seasonal ARIMA with exogenous variables (SARIMAX)

Different combinations of the components of the ARIMA class forecasting methodology were assayed using the statsmodels version 0.14.0 [4] and pmdarima auto-arima version 2.0.4 [5] packages. A seasonal ARIMA (0,0,[7])x(1,1,0,7) model, with exogenous variables (SARIMAX) was chosen as it offered the lower Akaike Information Criterion and higher Log-Likelihood [6] values. One-shot, seven-days ahead, 25 weekly predictions were built using all discharge observations until the eve of each index date for training, and assigning to the test period all observations from each index date up to six additional days ahead. Independent predictions were done for each department and ward using the predicted number of discharges (obtained with LGBM LOS predictions), non-working days and holiday eves as covariates.

##### Hospital discharge predictions using Prophet

One-shot, seven-days ahead, weekly discharge predictions were also generated with Facebook’s Prophet version 1.1.5 [7], using identical train-test splitting and covariates as for SARIMAX.

##### Hospital discharge predictions using LSTM

For LSTM predictions, a test period of 21 days preceding each index date and spanning 7 days into the future, was established. The validation phase covered 80 days prior to the test period, while the training data extended from 1^st^ January 2021 until the beginning of the validation period. One-day sliding window was used to generate supervised arrays with an input length of 21 days and an output length of 7 days, for the training, validation and test datasets. Scikit-learn [8] MinMaxScaler was used to scale all variables from −1 to +1 values. LSTM models were built with the TensorFlow package, using root mean squared error as the loss function. Training and validation loss plots were used to control overfitting. Final predictions were obtained after inverse transforming the scaled predictions on the test datasets.

##### Hospital discharge predictions using TFT

Pytorch-Forecasting version 1.0.0, TemporalFusionTransformer package [9] was used for TFT forecasts. Test datasets extended from 70 days before each index date until 7 days after this date. The validation dataset included the observations of the last 7 days before each test dataset, and the training datasets extended from 1^st^ January 2021 until the beginning of each validation dataset. One-day sliding window was used to generate supervised arrays with an input length of 70 days and an output length of 7 days. Early stopping on validation losses, patience limits, and validation loss plots with TensorBoard were used to control overfitting. Different attention head sizes, hidden sizes and learning rates were empirically tried to obtain the best results. Quantile loss was the metric used for the loss function.

##### Hospital discharge predictions using N_HiTS

Pytorch-Forecasting version 1.0.0 N_HiTS package was used for N_HiTS forecasts [9]. The same train, validation and tests timeseries dataframes and one-day sliding windows arrays used for TFT’s forecasting served for N_HiTS forecasts. Early stopping on validation losses, patience limits, and validation loss plots on TensorBoard were also used to control overfitting. MQF2DistributionLoss was the metric applied for the loss function.

##### Hospital discharge predictions using TCN

TensorFlow and Keras version 2.14.0 [10] packages were used to build TCN models. Independent models were created for overall hospital discharges and for each major hospital departments. In addition, weekly predictions with individual forecasts from day one until seven days ahead were generated using convolutional widths of 7 days and six stacked convolutional layers with progressive dilations. A validation dataset was used for each prediction. Validation losses, patience limits and validation loss plots on TensorBoard were used to control overfitting. MAE was the metric used for the loss function.

## 2.-Supplementary Tables

**Supplementary Table 1.** Impact of comorbidities on length of stay (multiple linear regression)

|  | B<br>coefficient | Sig (p). | Lower 0.95<br>CI | Upper 0.95<br>CI | Applied<br>factor<sup>1</sup> |
| --- | --- | --- | --- | --- | --- |
| Abdominal neoplasm | 1.08 | .000 | .69 | 1.47 | 1.1 |
| Acute kidney failure | 1.47 | .000 | .84 | 2.10 | 1.5 |
| Acute pancreatitis | 1.17 | .001 | .50 | 1.85 | 1.2 |
| Anemia | 1.17 | .000 | .84 | 1.50 | 1,2 |
| Asthenia | 5.81 | .001 | 2.53 | 9.09 | 5.8 |
| Ataxia | 2.87 | .001 | 1.23 | 4.53 | 2.9 |
| Atrioventricular block | 1.54 | .012 | .34 | 2.73 | 1.5 |
| Cerebral haemorrhage | 1.99 | .000 | .99 | 2.98 | 2 |
| Chronic kidney failure | 2.03 | .000 | 1.74 | 2.31 | 2 |
| CNS tumour | 3.52 | .000 | 1.66 | 5.38 | 3.5 |
| Constitucional syndrome | 2.98 | .000 | 1.59 | 4.38 | 3 |
| Cranial traumatism | 2.67 | .000 | 1.63 | 3.70 | 2.7 |
| Dorsal vertebral fracture | 3.00 | .000 | 2.20 | 3.80 | 3 |
| Dyspnea | 2.13 | .006 | .62 | 3.64 | 2.1 |
| Failure to thrive | 1.88 | .000 | .87 | 2.90 | 1.9 |
| Femur fracture | 2.20 | .000 | 1.09 | 3.32 | 2.2 |
| Hypo or hipernatremia | 2.17 | .000 | 1.18 | 3.17 | 2.2 |
| Ischemic ictus | 1.33 | .000 | .92 | 1.74 | 1.3 |
| Limb arterial ischemia | 2.03 | .000 | 1.15 | 2.91 | 2 |
| Liver cirrhosis | 2.94 | .000 | 2.38 | 3.50 | 3 |
| Lumbocystalgia | 1.13 | .000 | .58 | 1.68 | 1.1 |
| Lumbosacral fracture | 3.53 | .000 | 2.05 | 5.02 | 3.5 |
| Lung neoplasm | 2.01 | .000 | 1.35 | 2.68 | 2 |
| Metastatic neoplasm | 2.71 | .000 | 1.50 | 3.92 | 2.7 |
| Myelopathy | 9.83 | .000 | 7.45 | 12.21 | 10 |
| Osteosynthesis infection | 6.29 | .000 | 3.12 | 9.47 | 6.3 |
| Pancytopenia | 6.39 | .000 | 3.78 | 8.99 | 6.4 |
| Pleuritis | 1.87 | .004 | .60 | 3.15 | 1.9 |
| Polytraumatism | 3.47 | .002 | 1.30 | 5.63 | 3.5 |
| Pulmonary embolism | 1.38 | .002 | .51 | 2.24 | 1.4 |
| Spontaneous bacterial<br>peritonitis | 6.62 | .020 | 1.06 | 12.18 | 6.6 |

<sup>1</sup>Factor applied to build the comorbidity index. CI: Confidence interval.

**Supplementary Table 2.** Accuracy of weekly predictions of length of stay for episodes of hospitalization.

| Period of prediction | LOS prediction (LGBM) |  |  |  | LOS predictions (MLP) |  |  |
| --- | --- | --- | --- | --- | --- | --- | --- |
|  | N | MAE | MdAE | R | MAE | MdAE | R |
| March 6<sup>th</sup> – 12<sup>th</sup>, 2023 | 267 | 5.90 | 2.53 | 0.46 | 6.18 | 2.98 | 0.34 |
| March 13<sup>th</sup> – 19<sup>th</sup>, 2023 | 272 | 5.69 | 2.51 | 0.42 | 5.78 | 2.54 | 0.42 |
| March 20<sup>th</sup> – 26<sup>th</sup>, 2023 | 249 | 4.76 | 2.60 | 0.49 | 5.10 | 3.15 | 0.45 |
| March 27<sup>th</sup> – April 2<sup>nd</sup>, 2023 | 281 | 4.28 | 2.36 | 0.41 | 4.43 | 2.33 | 0.40 |
| April 3<sup>rd</sup> – 9<sup>th</sup>, 2023 | 262 | 4.93 | 2.53 | 0.41 | 5.07 | 2.50 | 0.33 |
| April 10<sup>th</sup> – 16<sup>th</sup>, 2023 | 275 | 6.18 | 2.91 | 0.44 | 6.46 | 3.29 | 0.33 |
| April 17<sup>th</sup> – 23<sup>th</sup>, 2023 | 291 | 4.65 | 2.33 | 0.54 | 4.59 | 2.15 | 0.51 |
| April 24<sup>th</sup> – 30<sup>th</sup>, 2023 | 289 | 4.27 | 2.37 | 0.48 | 4.27 | 2.37 | 0.43 |
| May 1<sup>st</sup> – 7<sup>th</sup>, 2023 | 327 | 5.95 | 2.65 | 0.39 | 5.61 | 2.59 | 0.38 |
| May 8<sup>th</sup> – 14<sup>th</sup>, 2023 | 269 | 4.37 | 2.32 | 0.54 | 4.56 | 2.16 | 0.37 |
| May 15<sup>th</sup> – 21<sup>th</sup>, 2023 | 280 | 4.51 | 2.44 | 0.56 | 4.86 | 2.62 | 0.44 |
| May 22<sup>nd</sup> – 28<sup>th</sup>, 2023 | 275 | 5.44 | 2.58 | 0.55 | 5.72 | 2.49 | 0.48 |
| May 29<sup>th</sup> – June 4<sup>th</sup>, 2023 | 274 | 4.67 | 2.51 | 0.54 | 5.02 | 2.59 | 0.47 |
| June 5<sup>th</sup> – 11<sup>th</sup>, 2023 | 284 | 5.16 | 2.39 | 0.53 | 5.30 | 2.35 | 0.45 |
| June 12<sup>th</sup> – 18<sup>th</sup>, 2023 | 269 | 4.43 | 2.22 | 0.47 | 4.72 | 2.23 | 0.42 |
| June 19<sup>th</sup> – 25<sup>th</sup>, 2023 | 282 | 4.31 | 2.33 | 0.48 | 4.57 | 2.30 | 0.39 |
| June 26<sup>th</sup> – July 2<sup>nd</sup>, 2023 | 291 | 3.89 | 2.31 | 0.56 | 4.37 | 2.39 | 0.46 |
| July 3<sup>rd</sup> – 9<sup>th</sup>, 2023 | 300 | 4.70 | 2.42 | 0.51 | 4.91 | 2.40 | 0.38 |
| July 10<sup>th</sup> – 16<sup>th</sup>, 2023 | 274 | 4.96 | 2.37 | 0.54 | 5.36 | 2.23 | 0.40 |
| July 17<sup>th</sup> – 23<sup>th</sup>, 2023 | 286 | 4.10 | 2.66 | 0.44 | 4.37 | 2.50 | 0.36 |
| July 24<sup>th</sup> – 30<sup>th</sup>, 2023 | 280 | 4.44 | 2.41 | 0.46 | 4.70 | 2.07 | 0.38 |
| July 31<sup>st</sup> – August 6<sup>th</sup>, 2023 | 257 | 4.54 | 2.48 | 0.50 | 5.14 | 2.85 | 0.38 |
| August 7<sup>th</sup> – 13<sup>th</sup>, 2023 | 234 | 3.94 | 2.19 | 0.57 | 4.12 | 2.05 | 0.48 |
| August 14<sup>th</sup> – 20<sup>th</sup>, 2023 | 254 | 4.30 | 2.62 | 0.45 | 4.33 | 2.59 | 0.41 |
| August 21<sup>st</sup> – 27<sup>th</sup>, 2023 | 227 | 3.83 | 2.34 | 0.51 | 3.72 | 2.23 | 0.52 |
| **Mean (SD)** | **274 (21)** | **4.7 (0.7)** | **2.5 (0.2)** | **0.5 (0.1)** | **4.9 (0.7)** | **2.5 (0.3)** | **0.4 (0.1)** |

LOS: length of hospital stay. LGBM: light gradient boosting machines. MLP: multilayer perceptron. MAE: mean absolute error. MdAE: median absolute error. R: Pearson's correlation. SD: standard deviation

**Supplementary Table 3.** Factors associated with higher errors in LOS predictions.

| Variables | MAE if present | MAE if absent | Correlation with MAE | Univariate analysis<sup>1</sup><br>*p*-value | Multivariate regression<br>*p*-value |
| --- | --- | --- | --- | --- | --- |
| *Categorical* |  |  |  |  |  |
| Male gender, mean (SD) | 5.6 (10.6) | 4.2 (8.0) |  | <0.001 | NS |
| Medical admission, mean (SD) | 6.2 (10.4) | 3.3 (7.8) |  | <0.001 | <0.001 |
| Surgical admission, mean (SD) | 5.7 (10.3) | 4.6 (9.1) |  | <0.001 | <0.001 |
| Use of B-S antibiotics, mean (SD) | 7.2 (11.7) | 4.7 (9.1) |  | <0.001 | <0.001 |
| *Continuous* |  |  |  |  |  |
| Age |  |  | 0.12 | <0.001 | NS |
| Comorbidity index |  |  | 0.11 | <0.001 | <0.001 |
| Mean LOS by main diagnosis |  |  | 0.29 | <0.001 | <0.001 |
| Mean LOS by diagnosis, PG |  |  | 0.25 | <0.001 | <0.001 |
| Mean LOS by diagnosis, OG |  |  | 0.24 | <0.001 | <0.001 |
| Median LOS by ward |  |  | 0.22 | <0.001 | <0.001 |
| Median LOS by service |  |  | 0.24 | <0.001 | <0.001 |
| Time in ED before admission |  |  | 0.11 | <0.001 | NS |

<sup>1</sup>t-test for the analysis of association between MAE values with categorical variables, Pearson's R for correlating MAE values with continuous variables; B-S antibiotics: broad-spectrum antibiotics; MAE: mean absolute error; LOS: length of stay; PG: pathophysiologically grouped; OG: organ-disease grouped. ED: Emergency department.

## 3.-Supplementary Figures

**Supplementary Figure 1.**
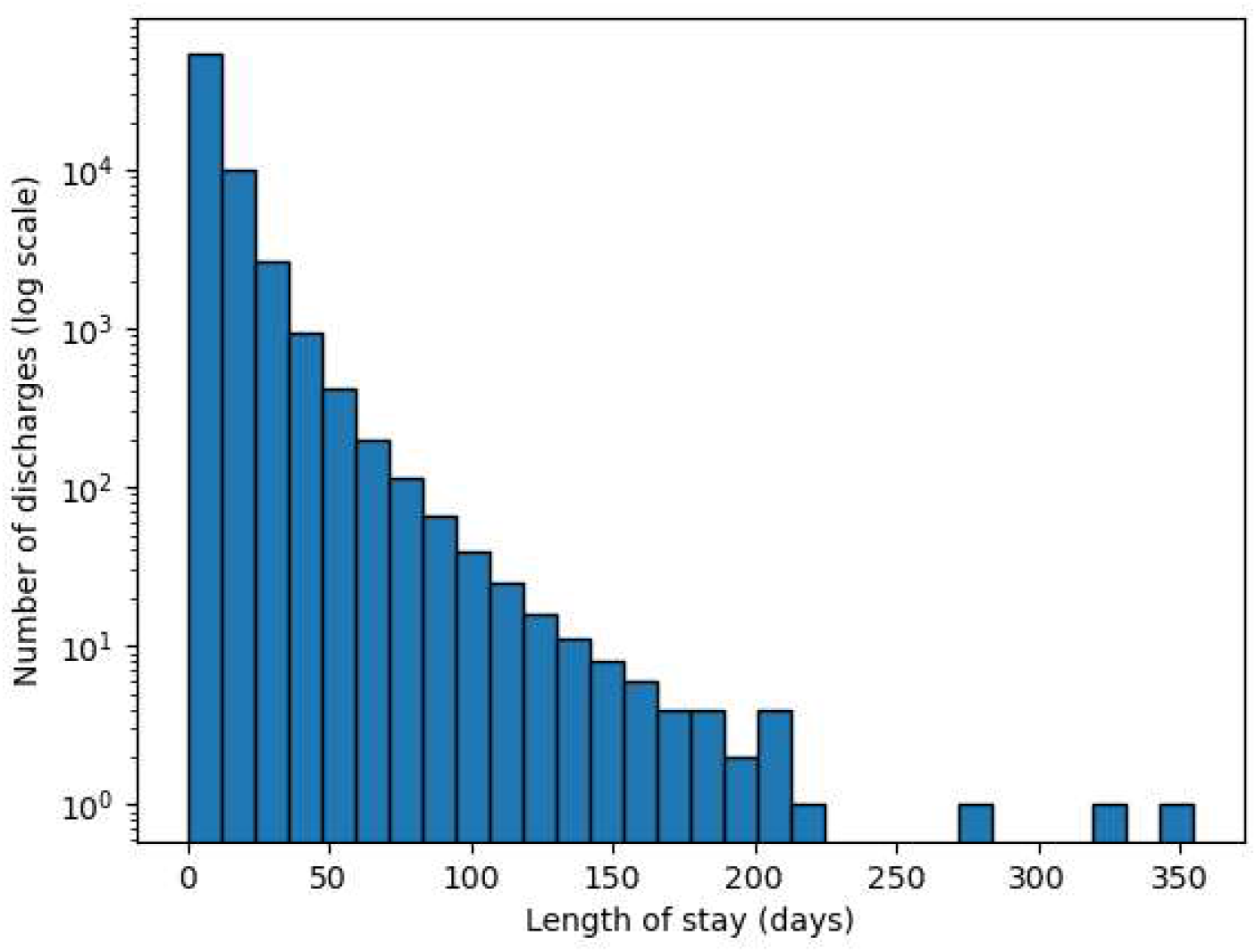
Distribution of length of stays in 67,308 hospitalization episodes proceeding from the Emergency department.

**Supplementary Figure 2.**
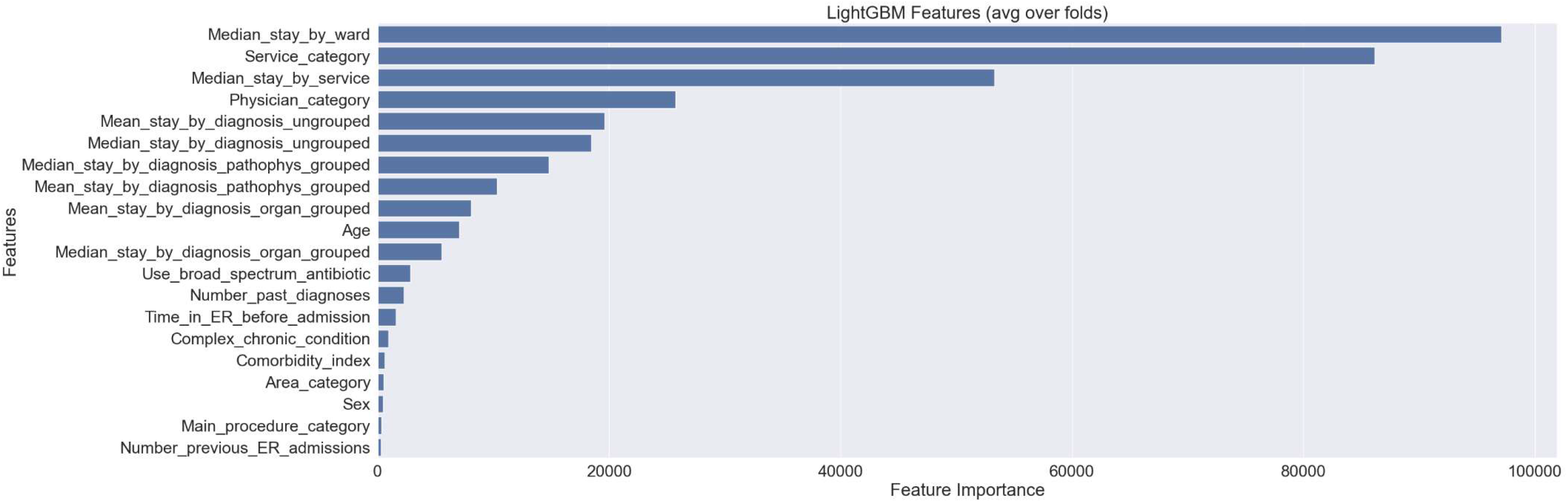
Feature importance in the prediction of length of hospital stay using Light Gradient Boosting Machines.

**Supplementary Figure 3.**
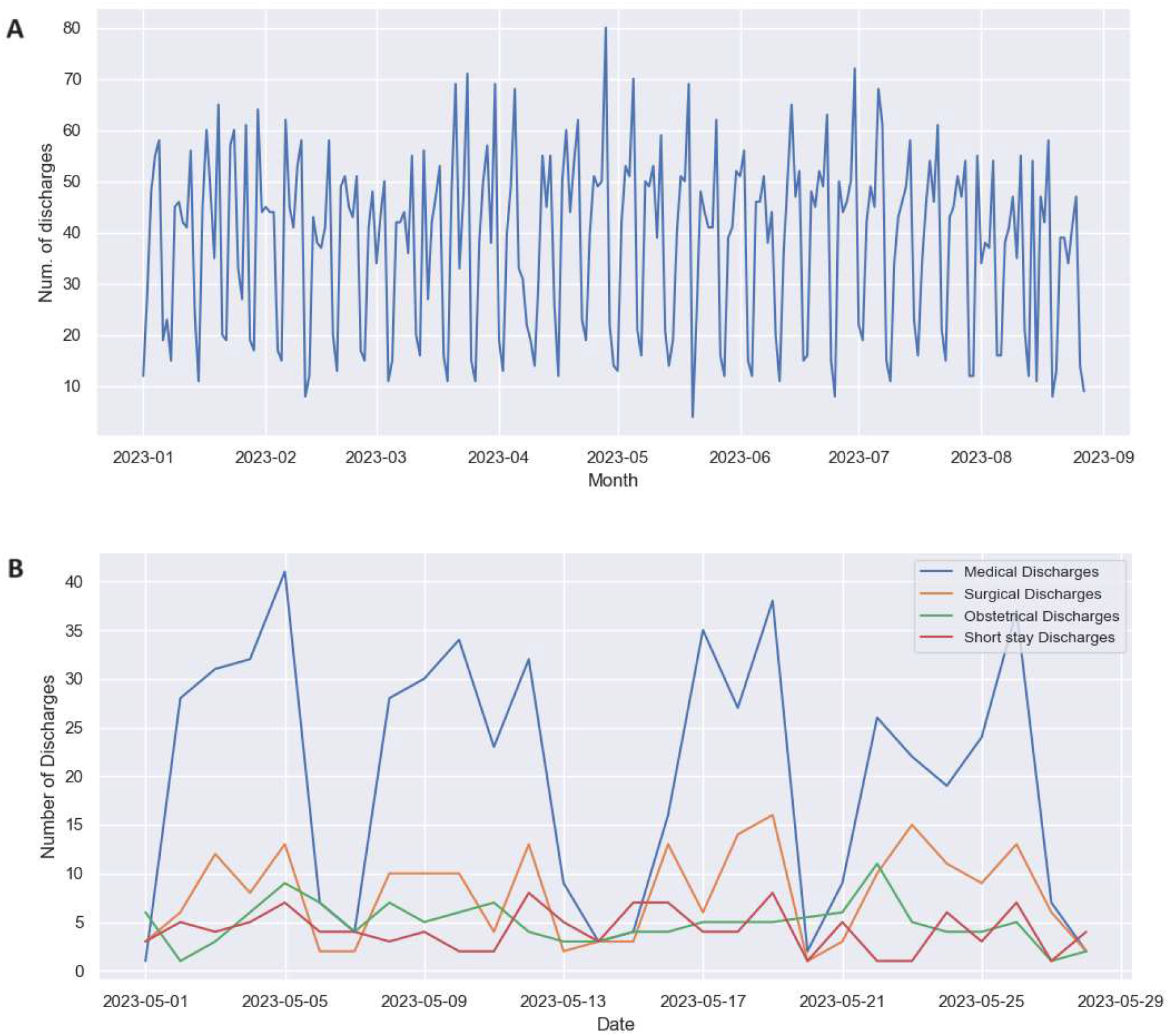
Seasonality of hospital discharges in patients admitted from the Emergency department. A: Overall discharges. B: Hospital discharges by department (sample May 2023).

**Supplemental Figure 4.**
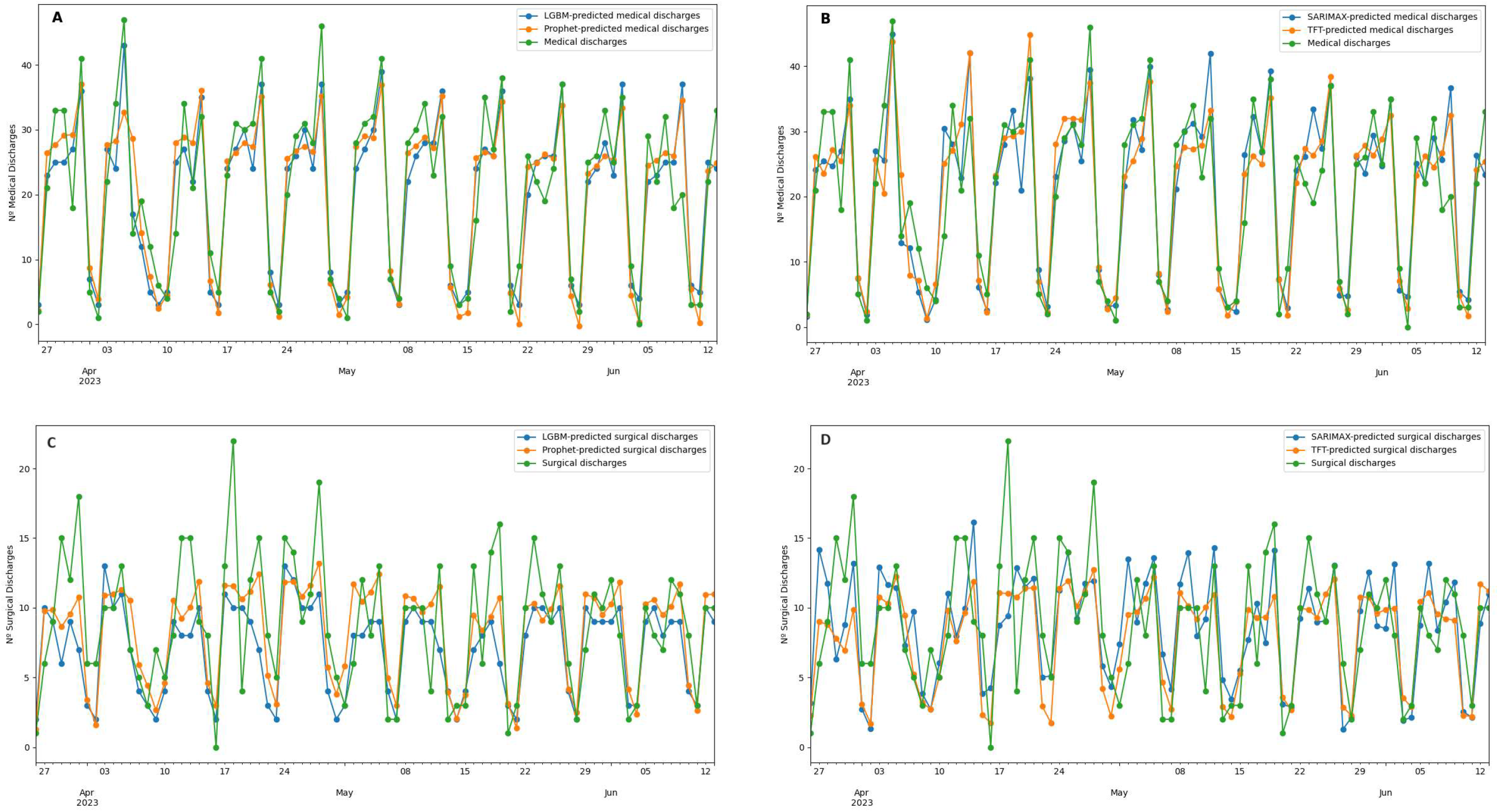
Concatenated weekly predictions of discharges. A: Light Gradient Boosting Machines and Prophet in Medical Deapartment. B: SARIMAX and Temporal Fusion Transformer in Medical Department. C: Light Gradient Boosting Machines and Prophet in Surgical Deapartment. B: SARIMAX and Temporal Fusion Transformer in Surgical Department.

**Supplemental Figure 5.**
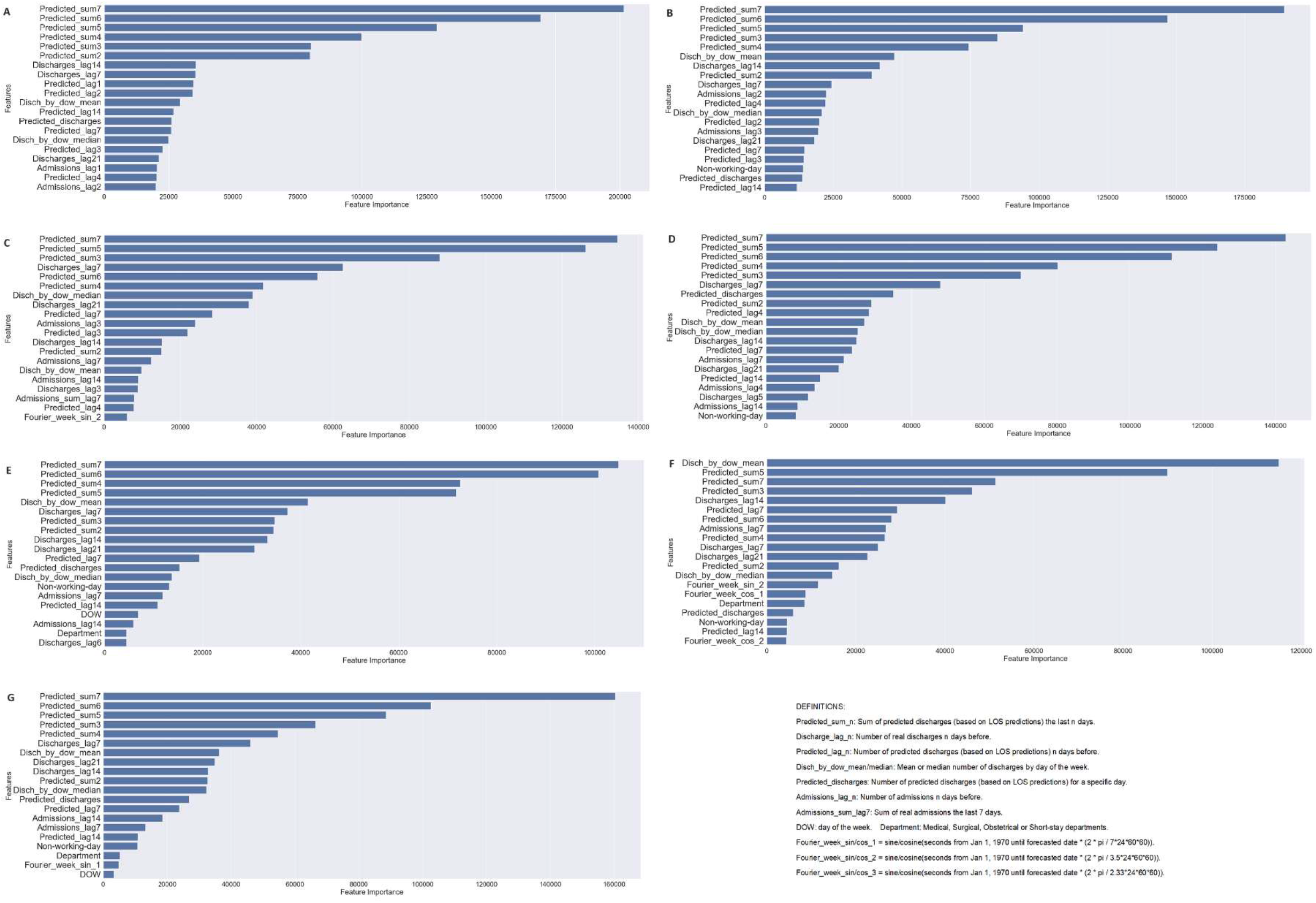
Feature importance for LGBM hospital discharge predictions: A: Model for first day prediction; B: Model for 2nd day prediction; C: Model for 3rd day prediction; D: Model for 4th day prediction; E: Model for 5th day prediction; F: Model for 6th day prediction; G: Model for 7th day prediction.

## ETHICAL APPROVAL

This study was approved by the ethics committee of the Corporació Sanitària Parc Taulí.

## ACKNOWLEDGEMENTS

Tomás Gil (Digital Strategy department, Corporació Sanitària Parc Taulí) for his participation in the acquisition of data.

Vicens Gaitán (SCALAI Scientific Director. Barcelona) for his contribution in the design of this study.

Ricard Comet (Internal Medicine Department, Corporació Sanitària Parc Taulí) and Xavier Bonfill (Epidemiology Department, Hospital de la Santa Creu I Sant Pau, Barcelona) for their useful comments on the manuscript.

